# Genome-wide association study of corneal dystrophy uncovers novel risk loci and enables improved polygenic prediction of Fuchs endothelial corneal dystrophy

**DOI:** 10.64898/2026.02.10.26345409

**Authors:** Benyapa Insawang, David A Mackey, Alex W Hewitt, Jamie E Craig, Richard Mills, Puya Gharahkhani, Stuart MacGregor

## Abstract

**Objective:** To identify risk loci for Fuchs endothelial corneal dystrophy (FECD) and improve a genetic risk prediction model.

**Design:** Genome–wide association study (GWAS), polygenic risk score (PRS) construction, and TCF4 CTG18.1 short tandem repeat (STR) length inference.

**Participants:** The study included 7,316 Europeans (EUR) with FECD or related corneal dystrophy phenotypes and 1,588,467 controls from the UK Biobank, All of Us, FinnGen, and the Million Veteran Program. Two independent EUR FECD cohorts were used for PRS validation (1,851/2,679 cases/controls and 124/257 cases/controls). African (AFR) ancestry analyses included 455 cases and 121,154 controls to build PRS. A subset of All of Us participants was used for joint PRS and STR modelling.

**Methods:** GWAS meta–analyses were performed using FECD diagnoses or corneal dystrophy proxies where necessary, with validity assessed via genetic correlation. Risk loci were identified, and ancestry–specific PRSs were constructed using SBayesRC. PRS performance was evaluated across ancestries with and without TCF4 STR data.

**Main Outcome:** We identified novel loci for corneal dystrophy and constructed PRS–based and STR–based prediction models.

**Results:** The GWAS meta–analysis identified 24 risk loci associated with corneal dystrophy, including 12 novel loci, doubling previous FECD studies. The optimised PRS outperformed existing models in two independent FECD validation cohorts (AUC = 0.83, 95% CI: 0.82–0.84; DeLong’s *P* = 7.04 × 10⁻^19^), with individuals in the top PRS decile showing 14–fold and 19–fold increased risk in the two validation sets, respectively

In All of Us, STR expansion (>40 repeats) was the key predictor of FECD risk, yielding excellent discrimination (AUC = 0.89; OR = 54) with minimal improvement from PRS. Consistent with this, STR expansion remained the primary driver of risk across ancestries, while PRS provided modest independent value for broader corneal dystrophy phenotypes in EUR and admixed American populations.

Among participants without large STR expansion, overall predictive performance was modest; PRS was the only significant genetic contributor (OR = 1.37) for broader corneal dystrophy in Europeans, whereas analyses in FECD non–expansion carriers were underpowered.

**Conclusions:** These findings refine the genetic architecture of FECD, enhance risk prediction, and support a tiered strategy integrating STR expansion testing with PRS.

**Key Points:** *Question:* Can polygenic risk scores (PRS), alone or combined with TCF4 CTG18.1 short tandem repeat (STR) length, improve genetic risk prediction for Fuchs endothelial corneal dystrophy (FECD)?

*Findings:* In this GWAS meta–analysis of 7,316 cases and 1,588,467 controls, PRS showed strong predictive performance in validation cohorts lacking STR data. When STR length was available, it was the main predictor of FECD risk with limited additional contribution from PRS. Among non–expansion STR carriers, PRS helped stratify risk for broader corneal dystrophy in Europeans.

*Meaning:* PRS provide a practical, complementary approach for FECD risk prediction, particularly when STR data are unavailable.

## Introduction

Fuchs’ endothelial corneal dystrophy (FECD) is a hereditary disease and the leading cause of corneal transplantation^1,2^. It is characterised by progressive endothelial dysfunction that complicates both diagnosis and treatment. FECD is a growing global health burden and contributes to rising healthcare costs in the United States^3–5^. Heritability estimates are approximately 30%^6^. Combining image analysis, genetic testing, and family history enables earlier FECD detection, helping prevent irreversible corneal damage^7^. FECD increases the risk of endothelial cell loss during cataract surgery, underscoring the need for preoperative endothelial assessment. When specular microscopy is unavailable, genetic testing, particularly a FECD polygenic risk score (PRS), may offer useful complementary risk stratification as genomic data become integrated into routine care.

Genetic risk prediction for FECD remains limited. Although GWAS have identified multiple susceptibility loci, most notably the transcription factor 4 (*TCF4*), and additional variants^8–10^, only one study has assessed FECD PRS, with modest predictive performance^9^. The CTG18.1 short tandem repeat (STR) expansion in *TCF4* strongly influences FECD risk and shows marked ancestry differences^11^, yet it has never been evaluated in a risk–prediction model or combined with PRS. Using whole–genome sequencing (WGS) data, we assessed the predictive value of both the STR expansion and PRS, enabling robust locus discovery and improved FECD risk prediction to support earlier detection and proactive clinical management.

## Methods

### Study Overview

We conducted a two–stage GWAS meta–analysis across four datasets: the UK Biobank (UKB)^12^, All of Us^13^, FinnGen (release 12)^14^, and the Million Veteran Program (MVP)^15^. From this, we created ancestry-specific PRSs for European and African populations. The first three datasets provided hereditary corneal dystrophy phenotypes, while MVP contributed FECD-specific data. The European PRS was further evaluated in two independent FECD cohorts. For STR analysis, we tested three PRSs in All of Us across the EUR, AMR, and AFR populations (Fig. 1).

**Figure 1:**
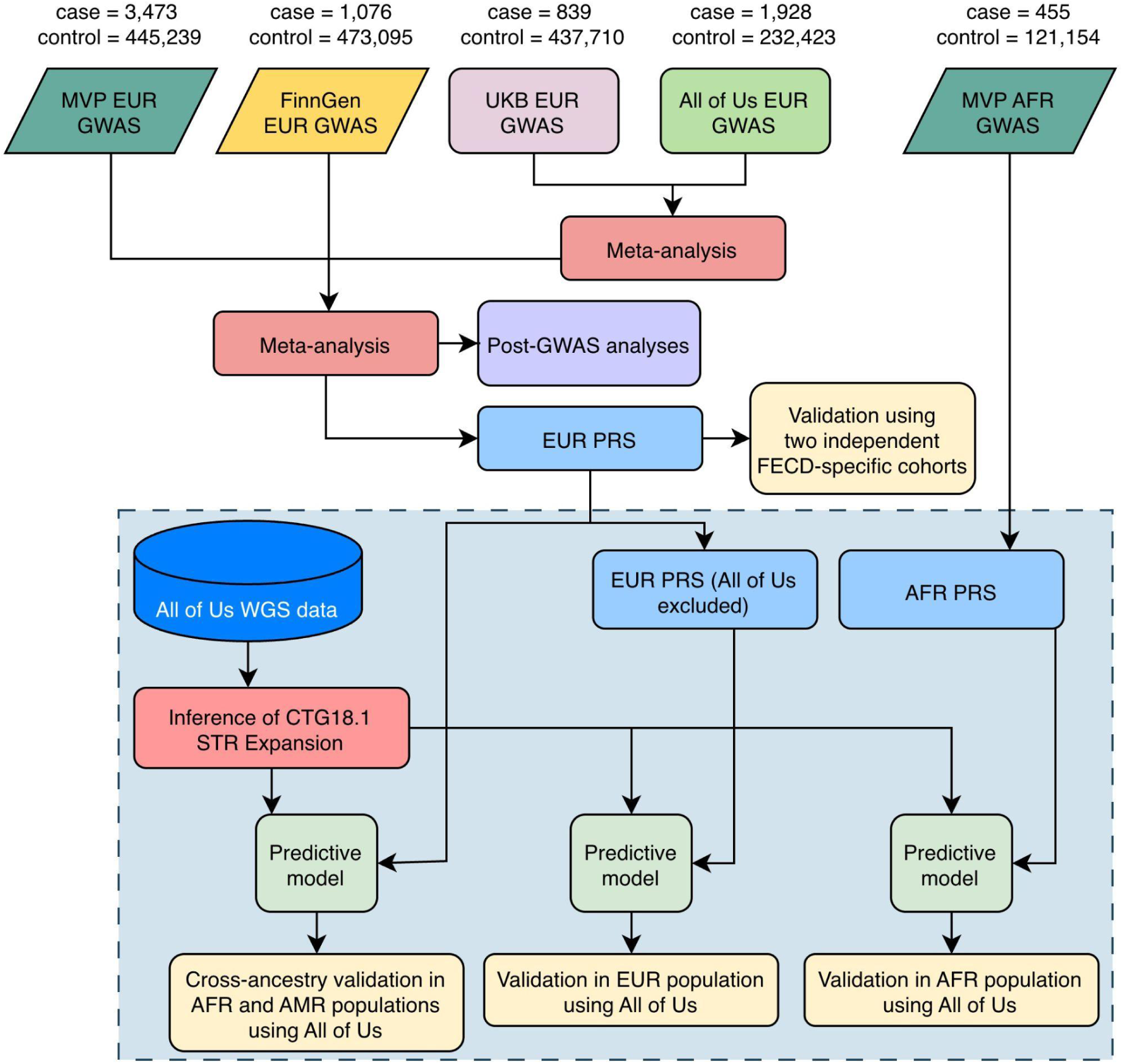
Study Overview. Genome-wide association study (GWAS) discovery analyses were conducted in the UK Biobank (UKB), All of Us, FinnGen, and the Million Veteran Program (MVP) among individuals of European (EUR) and African (AFR) ancestry. Meta-analyses were first performed using UKB and All of Us data, followed by a second-stage meta-analysis incorporating MVP and FinnGen. The number of Fuchs endothelial corneal dystrophy (FECD) cases and controls (or hereditary corneal dystrophy, where applicable) is shown. Polygenic risk scores (PRSs) were constructed separately for the EUR and AFR populations. The EUR PRS was evaluated in two independent EUR FECD cohorts, with array data available to compute a PRS but no sequencing data to accurately characterise the CTG18.1 short tandem repeat (STR). Three PRSs were used in the STR analyses: (i) the EUR PRS, evaluated in admixed American (AMR) and AFR populations; (ii) a recalculated EUR PRS excluding All of Us, evaluated in EUR individuals; and (iii) the AFR PRS, evaluated in AFR individuals. These PRSs were used to independently assess the predictive contributions of the STR expansion and PRS for FECD (or corneal endothelial dystrophy, where applicable).

**Discovery Dataset 1:** We identified 839 Europeans with hereditary corneal dystrophies (ICD-10: H185, Data-Field 41270) and 437,710 controls in UKB. GWAS was conducted with SAIGE (v1.3.0)^16^, adjusting for age, sex, and the first 10 principal components (PCs) of ancestry.

**Discovery Datasets 2:** In All of Us, 234,353 Europeans have WGS data. We performed a GWAS of hereditary corneal dystrophies with 1,928 cases identified by ICD-9 (371.5), ICD-10 (H18.5), or SNOMED (77797009), and 232,423 controls. Association testing used REGENIE (v3.2.6)^17^, adjusting for age, sex, and the first 10 PCs.

**Discovery Datasets 3:** The FinnGen GWAS studied hereditary corneal dystrophies (phenocode: H7_KERATOCONUS), with 1,076 cases and 473,095 controls of Finnish ancestry. The model included sex, age, 10 PCs, chip version, and genotyping batch as covariates in REGENIE. Although H7_KERATOCONUS implies keratoconus, it is classified under ICD-10 H18.5, and GWAS results do not indicate an enrichment for keratoconus-related single-nucleotide polymorphisms (SNPs).

**Discovery Datasets 4:** MVP conducted a GWAS on FECD (Phe_364_51) with 3,473 European cases and 445,239 controls. The MVP AFR cohort (455 cases, 121,154 controls) contributed to the AFR PRS. The GWAS was adjusted for age, sex, and the first 10 PCs using SAIGE.

**Validation Cohort 1:** We validated the PRS using two European datasets from dbGaP. The FECD GWAS (phs000421.v1.p1) contributed 1,856 FECD cases and 816 controls, and the AREDS Genetic

Variation in Refractive Error Substudy (phs000429.v1.p1) provided an additional 1,877 controls. Both studies were genotyped on the HumanOmni2.5 array. Quality control in PLINK (v1.9.0-b.7.7)^18^ removed variants with > 5% missingness, minor allele frequency (MAF) < 1%, Hardy–Weinberg P < 1×10⁻⁶, or samples with > 5% missing genotypes. Additionally, individuals with > 5% missingness on any chromosome were excluded, yielding a final dataset comprising 1,851 FECD cases and 2,679 controls. Genotypes were prepared using the HRC-1000G-checkbim tool (https://www.well.ox.ac.uk/wwrayner/tools/) and imputed via the Trans-Omics for Precision Medicine (TOPMed) r3 reference panel on the TOPMed Imputation Server (https://imputation.biodatacatalyst.nhlbi.nih.gov/), phasing with Eagle v2.4^19^ and imputation with Minimac4^20^. For PRS construction, we retained imputed variants with R² ≥ 0.3 and MAF ≥ 0.01.

**Validation Cohort 2:** The Fuchs’ Corneal Dystrophy Secondary GWAS (dbGaP phs000246.v2.p1; 127 FECD cases, 259 controls) was the second validation cohort^21^. Because ancestry labels were unavailable, we inferred ancestry using PCA with 1000 Genomes reference populations. Genotypes (hg18) were lifted to hg19 using LiftOver^22^, and variants with > 5% missingness, MAF < 1%, Hardy–Weinberg P < 1×10⁻⁶, or samples with > 5% missingness, were removed. The HRC–1000G–checkbim tool was applied, and linkage disequilibrium (LD) pruning (200–kb window, 50–kb step, r² ≤ 0.25) was used to generate SNPs for PCA. Ancestry was predicted using a random forest classifier trained on the first 20 principal components, implemented in R using the randomForest package^23^. After excluding three non–European samples (125 cases, 258 controls remaining), we applied the same QC pipeline as the first cohort for imputation.

**Validation Cohort 3.** WGS data from All of Us were used to infer the CTG18.1 STR expansion and assess its association with FECD, making this the third validation cohort. Because FECD case numbers were limited (80 European, 6 African, 1 South Asian), FECD–specific analyses were restricted to Europeans. To increase power and allow multi–ancestry comparisons, we used a broader corneal endothelial dystrophy (CED) phenotype, which yielded larger case counts: EUR (n = 935), AFR (n = 187), and AMR (n = 108). For each ancestry group, an equal number of age–matched controls (> 60 years) was selected to minimise confounding from age–related endothelial decline.

### Genome-Wide Association Study Meta-analysis

Because the four cohorts were generated using different study protocols, we first evaluated genetic correlations (r_g_) across them. The UKB dataset alone lacked power for stable r_g_ estimates, so we combined UKB and All of Us using an inverse–variance weighted fixed–effect meta–analysis in METAL (May 5, 2020)^24^ to improve power and harmonise phenotype mapping. Pairwise genetic correlations among the UKB/All of Us meta-analysis, FinnGen, and MVP were then estimated using LD Score Regression (LDSC)^25^. After confirming that all cohorts effectively captured the same underlying trait despite phenotyping differences, we performed a second inverse–variance fixed–effect meta-analysis that integrated summary statistics from UKB/All of Us, FinnGen, and MVP. Variants with MAF < 1% were removed prior to meta-analysis, and SNPs present in at least two datasets were submitted to FUMA (https://fuma.ctglab.nl/) for identification of independent genomic loci.

### Post-GWAS Analysis

We estimated SNP–based heritability (SNP–h^2^) from the GWAS meta–analysis using univariate LDSC, applying the baseline LD model and European reference LD scores from the 1000 Genomes Project. We conducted a phenome–wide association study (PheWAS) using newly identified SNPs, leveraging the MVP–FinnGen–UKB meta–analysis resource (https://mvp-ukbb.finngen.fi/) and Open Targets (https://platform.opentargets.org/). Gene–based analyses were performed using the mBAT–combo test implemented in GCTA (v1.94.1)^26^. We further conducted a transcriptome–wide association study (TWAS) using FUSION (February 1, 2022)^27^, integrating 1000 Genomes LD reference data and GTEx v8 whole–blood expression–weight panels (558 EUR samples) to identify genes whose genetically predicted expression is associated with FECD risk.

### PRS Validation

We constructed PRSs from the meta-analysis using SBayesRC, which estimates SNP effects while accounting for LD and functional annotations to improve predictive accuracy^26^. We compared our EUR PRS with two reference scores (a single *TCF4* SNP and a score using weights from the only published PRS for FECD to date^9^) and assessed whether adding the PRS improves performance beyond the traditional risk model.

Associations between each PRS and the FECD phenotype were evaluated using logistic regression. We normalised all PRSs, calculated the area under the receiver operating characteristic curve (AUC) from the linear predictors of the logistic regression models, and compared AUCs using DeLong’s test implemented in the pROC package^28^.

To quantify the contribution of major loci, we generated sensitivity PRSs by excluding variants within 1 Mb of *TCF4* or by removing chromosome 18 entirely, and evaluated changes in AUC and effect size. CTG18.1 STR expansion was inferred using ExpansionHunter (v5.0.0)^29^ on All of Us WGS data, and to isolate polygenic effects, the EUR PRS was recalculated, excluding All of Us when tested alongside STR results. PRS transferability was also assessed in non–European groups: an AFR–specific PRS was evaluated in AFR individuals, and the EUR PRS (trained on 4 cohorts) was tested in AFR and AMR populations.

## Results

### Genome-Wide Association Study Meta-analysis

The input datasets included FECD–defined cases (MVP) or hereditary corneal dystrophy cases (UKB, All of Us, and FinnGen). Despite differences in case definitions, pairwise genetic correlations were consistently high (r_g_ > 0.8, Table 1), confirming that the cohorts captured the same underlying genetic trait. After removing the *TCF4* region (±1 Mb), the MVP–FinnGen correlation remained high (r_g_ = 0.93, SE = 0.29), indicating the shared signal was not driven solely by *TCF4*.

**Table 1.** Pairwise genetic correlations (r_g_) between cohorts estimated using Linkage Disequilibrium Score Regression (LDSC).

| Cohort 1 | Cohort 2 | $r_g$ (SE) | P-value |
| --- | --- | --- | --- |
| FinnGen | Meta-analysed UKB/All of Us | 0.8902 (0.2138) | $3.12 \times 10^{-5}$ |
| FinnGen | MVP | 0.9516 (0.1464) | $8.10 \times 10^{-11}$ |
| Meta-analysed UKB/All of Us | MVP | 1.1336 (0.2878) | $8.18 \times 10^{-5}$ |
| FinnGen (removed *TCF4* region) | MVP (removed *TCF4* region) | 0.9327 (0.2886) | 0.0012 |

$r_g$ = genetic correlation; SE = standard error. P-values test the null hypothesis that $r_g = 0$ . Genetic correlations were high across cohorts, showing strong shared genetic architecture despite differing case definitions.

A meta-analysis of all cohorts identified 23 independent genome–wide significant SNPs (Table 2). Eleven of these, near or within *NID1*, *AKAP7*, *XYLT1*, *GRM5*, *NTMT2*, *DIO3*, *COL24A1*, *PPARGC1B*, *TNS3*, *RUNX2*, and *TUBA3C*, represent novel FECD–associated loci (P < 5 × 10⁻^8^). The strongest association was observed at *TCF4*, with rs11659764 showing an OR of 6.19 (95% CI: 5.87–6.53). Effect estimates for our findings were consistent with the latest FECD GWAS meta–analysis by Gorman et al.^10^. The SNP–h^2^ was estimated at 0.22 (SE = 0.13), assuming a 1% population prevalence.

**Table 2.** Genome-wide significant loci in the meta-analysis of corneal dystrophy in European ancestry.

| Nov<br>el | SNP | CH<br>R | POS | EA | NEA | EAF | OR | 95% CI | P | Directi<br>on |
| --- | --- | --- | --- | --- | --- | --- | --- | --- | --- | --- |
| 1 | rs1466901<br>75 | 1 | 23622855<br>7 | T | C | 0.03 | 1.46 | [1.31,<br>1.62] | $2.41 \times 10^{-12}$ | +++ |
| 1 | rs4895918 | 6 | 13144919<br>5 | T | C | 0.29 | 1.13 | [1.09,<br>1.17] | $3.23 \times 10^{-11}$ | +++ |
| 1 | rs1128838<br>3 | 16 | 17646392 | A | AC | 0.14 | 1.18 | [1.12,<br>1.24] | $8.64 \times 10^{-11}$ | +++ |
| 1 | rs1101843<br>9 | 11 | 88775561 | T | C | 0.31 | 1.13 | [1.09,<br>1.17] | $4.08 \times 10^{-10}$ | +++ |
| 1 | rs1015751<br>8 | 1 | 17017069<br>5 | A | C | 0.38 | 1.11 | [1.07,<br>1.15] | $1.53 \times 10^{-9}$ | +++ |
| 1 | rs1013979<br>7 | 14 | 10170860<br>9 | A | G | 0.14 | 1.15 | [1.10,<br>1.21] | $1.58 \times 10^{-9}$ | +++ |
| 1 | rs1336050 | 1 | 86688478 | T | G | 0.81 | 0.88 | [0.84,<br>0.92] | $3.40 \times 10^{-9}$ | --- |
| 1 | rs4705363 | 5 | 14906222<br>2 | A | G | 0.19 | 1.13 | [1.09,<br>1.18] | $6.57 \times 10^{-9}$ | +++ |
| 1 | rs700755 | 7 | 47445641 | A | G | 0.49 | 0.91 | [0.88,<br>0.94] | $7.14 \times 10^{-9}$ | --- |
| 1 | rs1934328 | 6 | 45465753 | A | T | 0.53 | 1.10 | [1.06,<br>1.14] | $1.84 \times 10^{-8}$ | +++ |
| 1 | rs1508286<br>50 | 13 | 19588831 | T | C | 0.02 | 1.41 | [1.25,<br>1.60] | $3.61 \times 10^{-8}$ | +++ |
| 0 | rs1388345<br>42 | 18 | 52788802 | A | G | 0.03 | 4.78 | [4.47,<br>5.11] | $4.30 \times 10^{-449}$ | +++ |
| 0 | rs1200108 | 1 | 16905796<br>9 | A | G | 0.65 | 0.78 | [0.75,<br>0.80] | $6.58 \times 10^{-49}$ | --- |
| 0 | rs7974289<br>5 | 1 | 62782860 | T | C | 0.95 | 0.64 | [0.59,<br>0.69] | $5.34 \times 10^{-31}$ | --- |
| 0 | rs8009540<br>9 | 7 | 10760021<br>1 | C | G | 0.02 | 1.67 | [1.49,<br>1.87] | $7.27 \times 10^{-19}$ | +++ |
| 0 | rs2333620 | 1 | 18310055<br>5 | T | C | 0.44 | 1.16 | [1.12,<br>1.19] | $6.05 \times 10^{-18}$ | +++ |
| 0 | rs1140658<br>56 | 21 | 46852758 | T | C | 0.04 | 0.64 | [0.58,<br>0.71] | $7.44 \times 10^{-18}$ | --- |
| 0 | rs1159055<br>7 | 1 | 54789772 | A | G | 0.04 | 1.46 | [1.33,<br>1.59] | $3.03 \times 10^{-17}$ | +++ |
| 0 | rs4963153 | 11 | 791462 | A | G | 0.48 | 1.15 | [1.12,<br>1.19] | $4.18 \times 10^{-17}$ | +++ |
| 0 | rs4138804<br>8 | 7 | 11729482 | A | C | 0.05 | 1.32 | [1.23,<br>1.42] | $7.89 \times 10^{-14}$ | +++ |
| 0 | rs1412082<br>02 | 20 | 60897104 | T | C | 0.04 | 1.34 | [1.23,<br>1.47] | $8.24 \times 10^{-11}$ | +++ |
| 0 | rs3515089<br>3 | 17 | 14549826 | A | AT | 0.37 | 0.87 | [0.83,<br>0.91] | $4.88 \times 10^{-10}$ | --? |
| 0 | rs1720438<br>1 | 15 | 61043150 | A | G | 0.92 | 0.84 | [0.79,<br>0.89] | $1.52 \times 10^{-9}$ | --- |

Genomic risk loci identified by FUMA from the meta-analysis of European ancestry cohorts (UK Biobank, All of Us, FinnGen, and MVP). FUMA detected 23 independent loci, including 11 novel corneal dystrophy loci and all 12 previously reported FECD loci. Genomic coordinates are based on the GRCh38 reference genome. Novel indicates whether the locus represents a newly identified association (1 = yes, 0 = no). EA: effect allele; NEA: non-effect allele; EAF: effect allele frequency; OR: odds ratio; 95% CI: confidence interval bounds. Direction denotes the effect direction in MVP, FinnGen, and the meta-analysed UKB/All of Us datasets, respectively; “?” indicates that the variant was unavailable in a given cohort.

### Comparative Performance of Polygenic Risk Scores (PRSs)

We assessed PRS performance in two independent validation cohorts: 4,530 European–ancestry individuals in the first cohort and 381 in the second. To evaluate predictive gains, we compared our new FECD PRS (PRS_2025_) with two reference scores: PRS_2017_, built from the top SNPs in the four replicated loci reported by Afshari et al.^9^, and PRS_2010_, a single–variant score at the *TCF4* locus using rs618869 as a proxy for rs613872 (R^2^ = 0.83, D′ = 1). The effect weight for rs618869 was derived using SBayesRC from our meta–analysis (OR = 2.26, PIP = 1), indicating high confidence in its contribution to disease risk.

The AUC for PRS_2025_ alone was 0.8314 (95% CI: 0.8188–0.8440), indicating strong predictive power, with high–PRS individuals at markedly elevated FECD risk (Table 3). PRS_2025_ significantly outperformed both comparison scores, PRS_2017_ (P = 7.04 × 10^-19^) and PRS_2010_ (P = 8.52 × 10^−63^), highlighting substantial improvement in risk prediction. Individuals in the top 10% of the PRS distribution had ORs of 19.04 (95% CI: 7.87–56.88) in cohort 2 and 13.97 (95% CI: 10.49–18.99) in cohort 1.

**Table 3.** Comparison of Predictive Performance Across Polygenic Risk Scores.

| Polygenic Risk Score | Odds Ratio per Standard Deviation (95% Confidence Interval) | Odds ratio per Standard Deviation comparing individuals in the top 10% of PRS versus the remaining (95% Confidence Interval) | Area under the Receiver Operating Characteristic Curve (95% Confidence Interval) | P Values (Areas under the Receiver Operating Characteristic Curve vs. PRS<sub>2025</sub>) |
| --- | --- | --- | --- | --- |
| **Validation Cohort 1** |  |  |  |  |
| PRS<sub>2025</sub> | 4.97 (4.51–5.49) | 13.97 (10.49–18.99) | 0.8314 (0.8188–0.8440) |  |
| PRS<sub>2017</sub> | 3.10 (2.87–3.35) | 5.31 (4.26–6.67) | 0.7756 (0.7618–0.7895) | $7.04 \times 10^{-19}$ |
| PRS<sub>2010</sub> | 2.29 (2.14–2.45) | 3.38 (2.76–4.16) | 0.6949 (0.6792–0.7107) | $8.52 \times 10^{-63}$ |
| **Validation Cohort 2** |  |  |  |  |
| PRS<sub>2025</sub> | 3.52 (2.61–4.96) | 19.04 (7.87–56.88) | 0.7819 (0.7317–0.8320) |  |
| PRS<sub>2017</sub> | 2.21 (1.76–2.82) | 4.38 (2.22–8.99) | 0.7039 (0.6464–0.7615) | $4.97 \times 10^{-3}$ |
| PRS<sub>2010</sub> | 1.70 (1.37–2.12) | 3.21 (2.04–5.08) | 0.6321 (0.5804–0.6838) | $6.52 \times 10^{-7}$ |

PRS<sub>2025</sub>: Polygenic risk score developed in this study; PRS<sub>2017</sub>: Score based on Afshar et al. GWAS summary statistics; PRS<sub>2010</sub>: Single-variant score at *TCF4* using rs618869 as a proxy for rs613872.

To assess added clinical value, we compared a baseline model (age, sex, top 10 PCs) with the same model including PRS_2025_. In cohort 1, AUC increased from 0.6157 (95% CI: 0.5992–0.6322) to 0.8468 (95% CI: 0.8351–0.8586). In cohort 2, AUC rose from 0.5784 (95% CI: 0.5164–0.6405) to 0.7966 (95% CI: 0.7467–0.8464), demonstrating substantial improvement beyond the traditional risk model. Excluding the *TCF4* region (±1 Mb) reduced PRS_2025_ performance from an AUC of 0.8468 to 0.7371 and lowered the effect size from OR = 5.16 to OR = 2.46. Removing all chromosome 18 variants further reduced AUC to 0.7157 (OR = 2.06), confirming the dominant influence of *TCF4* and nearby signals.

To separate the effect of the *TCF4* tag SNP from broader polygenic risk, we jointly modelled PRS_2025_ and rs613872. The combined model achieved the highest performance (AUC = 0.8546, 95% CI: 0.8432–0.8659), with both predictors remaining significant (PRS_2025_ OR = 3.83; rs613872 OR = 2.00). Removing the *TCF4* region from PRS_2025_ lowered AUC to 0.8236 and reduced the PRS effect (OR = 2.00), while increasing the rs613872 effect (OR = 4.82). Validation cohort 2 showed the same pattern, though with wider confidence intervals due to its smaller sample size.

### Prediction Model with Transcription Factor 4 (*TCF4*) CTG18.1 Short Tandem Repeat Length on Fuchs Endothelial Corneal Dystrophy (FECD)

Using WGS data from All of Us, we inferred STR expansion status (>40 repeats) and observed the expected ancestry pattern, with the highest carrier rate in EUR participants (Table 4). STR expansion status was the dominant predictor in the FECD predictive model, alone achieving the strongest discrimination (AUC = 0.8939; 95% CI, 0.8455–0.9423) and a very large effect size (OR = 54.21; 95% CI, 16.59–233.11). Adding the EUR PRS or the *TCF4* tag SNP rs613872 did not improve performance (ΔAUC ≤ 0.002), and neither was significant (P > 0.6).

**Table 4.** Summary of CTG18.1 Repeat Length and Demographic Characteristics by Corneal Endothelial Dystrophy (CED) or Fuchs Endothelial Corneal Dystrophy (FECD) Status.

| Ancestry | Disease Status | Mean | Median | Min | Q1 | Q3 | Max | N | Mean Age | N Male | N Female | Expanded (N, %) |
| --- | --- | --- | --- | --- | --- | --- | --- | --- | --- | --- | --- | --- |
| **Corneal Endothelial Dystrophy** |  |  |  |  |  |  |  |  |  |  |  |  |
| **African** | Control | 22 ± 11 | 21 | 11 | 17 | 24 | 120 | 187 | 69 ± 6 | 72 | 115 | 4 (2.14) |
|  | Case | 32 ± 26 | 22 | 11 | 17 | 26 | 133 | 187 | 70 ± 11 | 32 | 155 | 38 (20.32) |
| **European** | Control | 24 ± 18 | 17 | 11 | 15 | 25 | 118 | 935 | 73 ± 7 | 387 | 548 | 74 (7.91) |
|  | Case | 50 ± 34 | 38 | 11 | 17 | 79 | 153 | 935 | 77 ± 9 | 388 | 547 | 467 (49.95) |
| **Admixed American** | Control | 21 ± 15 | 17 | 11 | 15 | 19 | 95 | 108 | 70 ± 7 | 38 | 70 | 6 (5.56) |
|  | Case | 36 ± 28 | 23 | 11 | 17 | 62 | 129 | 108 | 69 ± 12 | 33 | 75 | 31 (28.70) |
| **Fuchs Endothelial Corneal Dystrophy** |  |  |  |  |  |  |  |  |  |  |  |  |
| **European** | Control | 26 ± 23 | 17 | 11 | 14 | 25 | 131 | 80 | 72 ± 9 | 36 | 44 | 8 (10.00) |
|  | Case | 60 ± 32 | 67 | 11 | 25 | 83 | 135 | 80 | 75 ± 8 | 23 | 57 | 54 (67.50) |

Summary statistics for CTG18.1 short tandem repeat (STR) length and demographic characteristics across ancestries and case–control status for corneal endothelial dystrophy (CED) and Fuchs endothelial corneal dystrophy (FECD). STR length metrics include mean ± SD, median, quartiles (Q1–Q3), and range (min–max) based on the maximum allele per individual. Expanded (N, %) indicates the number and proportion of individuals with STR expansion defined as >40 repeats.

### Prediction Model with Transcription Factor 4 (*TCF4*) CTG18.1 Short Tandem Repeat Length on Corneal Endothelial Dystrophy (CED)

For AFR and AMR ancestries, insufficient FECD cases in All of Us precluded direct validation. Instead, we used the closest available phenotype, CED.

Among AFR participants (n = 374), STR expansion status emerged as the primary driver of prediction, offering higher discrimination (AUC = 0.7325, 95% CI: 0.6825–0.7825; OR = 14.77, 95% CI: 5.42–52.44) than the AFR PRS (AUC = 0.6882, 95% CI: 0.6348–0.7415; OR = 1.37, 95% CI: 1.12–1.73). Once STR status was included, the PRS lost significance, a pattern that persisted when the EUR PRS was applied.

In contrast, AMR individuals (n = 216) showed strong cross–ancestry PRS performance: the EUR PRS substantially improved prediction (AUC = 0.7907, 95% CI: 0.7310–0.8505; OR = 2.00, 95% CI: 1.46–2.90), while STR expansion status offered lower predictive value (AUC = 0.7706, 95% CI: 0.7084–0.832; OR = 6.36, 95% CI: 2.42–19.33). In the combined model, the PRS remained significant (OR = 1.77, 95% CI: 1.19–2.75, P = 0.007), whereas the STR effect was not statistically significant, yielding an AUC of 0.7876 (95% CI: 0.7273–0.8479).

In EUR individuals, the recalculated EUR PRS (excluding All of Us) improved prediction over baseline from 0.6252 (95% CI: 0.5999–0.6504) to 0.7474 (95% CI:0.7253–0.7695), while STR expansion status provided the strongest single–predictor performance (AUC = 0.7782, 95% CI: 0.7567–0.7996; OR = 12.04, 95% CI: 9.19–15.96). In the combined model, PRS remained independently associated (OR = 1.30, 95% CI: 1.16–1.46), offering only marginal improvement beyond expansion status alone.

### Prediction Model in Non-expansion Carriers

Because many individuals with FECD or CED do not carry large STR expansions, we assessed whether PRS adds value in STR non–expansion carriers. Across ancestries, predictive ability was modest. In EUR CED non–expansion carriers (468 cases, 861 controls), PRS provided the only significant genetic effect (OR = 1.37, 95% CI: 1.17–1.60, P = 1.20×10⁻⁴) and yielded small AUC gains over baseline. PRS similarly increased AUC in AFR and AMR groups, though not significantly. Analysis of FECD non–expansion carriers was underpowered (26 cases, 73 controls); PRS improved discrimination over demographic covariates alone but was not statistically significant.

### Gene-Based and Transcriptome-Wide Association Study (TWAS) Results

We mapped meta–analysis SNPs to 17,648 genes for gene-based testing and identified 34 surpassing Bonferroni significance (P = 2.83 × 10⁻⁶), but no additional loci emerged beyond those detected in the SNP–level meta–analysis. A TWAS identified a single Bonferroni–significant gene, *WWP2* (rs3748387; P < 3.04 × 10^−6^). Locus-level colocalization was not performed because eQTL data from corneal tissues are currently unavailable. Consequently, *WWP2* should be considered a provisional candidate, pending colocalization and replication analyses in corneal datasets.

### Phenome-Wide Association Study (PheWAS)

We examined cross–trait associations for the 12 newly identified SNPs. Six variants (rs4895918, rs4705363, rs700755, rs1934328, rs15082865, and rs3748387) showed non–significant associations with hereditary corneal dystrophy in the MVP–FinnGen–UK Biobank meta-analysis platform. Open Targets recorded that five SNPs (rs11288383, rs11018439, rs10157518, rs10139797, and rs1336050) were linked to eye–related traits, whereas rs700755 showed associations with skin cancer phenotypes and rs3748387 with tobacco smoking–related traits.

## Discussions

We conducted the largest GWAS meta-analysis of corneal dystrophy to date, incorporating 7,316 cases and 1,588,467 controls across four cohorts^12–15^. By analysing FECD jointly with hereditary corneal dystrophies, we identified 12 previously unreported loci, doubling the number of known FECD risk loci from 12 to 24 (23 from the meta-analysis and one from the TWAS). These findings enabled the construction of an improved PRS that outperformed prior models. Our PheWAS further supported the associations observed in earlier GWAS of corneal dystrophy.

Among the novel loci, *XYLT1*, *COL24A1*, and *NID1* stand out for their roles in extracellular matrix and basement membrane structure, which aligns with known FECD pathology. We also identified *GRM5* and *WWP2*, genes previously implicated in ocular structural traits and endothelial regulation^30,31^, further supporting their potential relevance to FECD.

This study has several limitations related to phenotype definition and cohort composition. In some cohorts, FECD was not directly recorded, so hereditary corneal dystrophy was used as a proxy, which may have introduced heterogeneity despite high genetic correlations across datasets. Given these correlations, we used inverse–variance–weighted meta–analysis (METAL) rather than MTAG^32^, as METAL performs comparably under high trait correlation and retains more SNPs for downstream analyses. Additionally, the lower observed FECD prevalence compared with global estimates likely reflects underdiagnosis and incomplete clinical ascertainment.

We constructed our FECD PRS using SBayesRC, incorporating 6.89 million variants to reflect the disorder’s highly polygenic architecture. This represents a major advance over earlier scores, PRS_2010_ (1 SNP) and PRS_2017_ (4 SNPs), and explains the markedly stronger associations achieved by our model (PRS_2017_: P = 7.04 × 10^-19^; PRS_2010_: P = 8.52 × 10^−63^). Performance trends were consistent across validation cohorts, underscoring the distributed nature of FECD genetic risk.

Combining PRS2025 (with the *TCF4* region removed) with rs613872 improved prediction (AUC = 0.8236 vs. 0.7942 for rs613872 alone). Although collinearity indicates that *TCF4* substantially contributes to the PRS signal, removing this region reduced the PRS effect and increased the effect of rs613872, showing that the PRS captures additional genome–wide risk beyond *TCF4*. These results demonstrate that the broader polygenic background provides meaningful independent predictive value and enables more refined risk stratification than single–variant models.

The strong performance of our PRS has important clinical implications. By leveraging nearly seven million variants, it captures FECD’s polygenic risk more fully and enables early identification of high–risk individuals before clinical signs appear. Because FECD increases the likelihood of endothelial decompensation after cataract surgery, the PRS could complement pre–operative assessment and improve surgical planning. It may also aid glaucoma management, particularly when considering procedures such as drainage device implantation that can accelerate endothelial loss. More broadly, the PRS supports personalised monitoring for mid–life individuals without visible guttae but with elevated genetic risk.

Our findings showed that CTG18.1 STR expansion status remains the strongest single predictor of FECD risk (AUC = 0.89; OR = 54), although its contribution varies across ancestries within the broader phenotype, as few FECD cases are available outside of EUR. In EUR and AFR individuals, expansion status dominated risk prediction, whereas in AMR individuals, the PRS retained significant predictive value even after accounting for STR status. Importantly, in the EUR CED non–expansion carriers, who represent a clinically challenging subgroup, the PRS provided meaningful additional discrimination. However, analyses of FECD non–expansion carriers were underpowered, and larger cohorts with STR data are needed to clarify whether PRS improves risk stratification in this subgroup.

These findings support a tiered genetic risk–prediction framework: prioritise *TCF4* STR expansion testing where feasible, use PRS when expansion data are unavailable, and combine both for optimal performance. Because most large biobanks rely on genotyping arrays rather than WGS, PRS represents a practical, scalable tool that can be generated from existing data without specialised STR assays. Additionally, given that expansion length correlates with severe, late–stage FECD, PRS may help identify individuals at risk for earlier or milder disease, enabling earlier monitoring and intervention.

In summary, we identified 12 novel loci, substantially expanding the known genetic architecture of FECD. Building on these discoveries, we developed a genome–wide PRS that achieved markedly superior performance compared with previous SNP–based models. Our findings highlight the power of integrating polygenic signals at scale while reaffirming the strong predictive value of the *TCF4* STR expansion, which should remain the primary genetic assay when available. Together, *TCF4* expansion testing and advanced PRS offer complementary, scalable, and clinically actionable tools for improving genetic risk assessment and provide a foundation for future personalised screening and early–intervention strategies.

## Data Availability

All data produced in the present study are available for non-commercial use upon reasonable request to the authors.

## Acknowledgments

Funding/Support

SM acknowledges Program Grant (1150144) and Fellowship (2034568) funding from the Australian National Health and Medical Research Council (NHMRC). PG acknowledges NHMRC Investigator Grant (1173390). AWH and JEC are supported by NHMRC Investigator grants. DAM is supported by NHMRC, Glaucoma Australia, and Telethon.

## Other Acknowledgments

This research has been conducted using data from the UK Biobank (application number 25331). We thank the participants and investigators of UK Biobank, Finngen, and the Million Veteran Program.

We gratefully acknowledge All of Us participants for their contributions, without whom this research would not have been possible. We also thank the National Institutes of Health’s All of Us Research Program for making available the participant data examined in this study.

The dataset(s) used for the analyses described in this manuscript were obtained from the Genetics of Fuchs’ Endothelial Corneal Dystrophy (FECD) Study found at https://www.ncbi.nlm.nih.gov/projects/gap/cgi-bin/study.cgi?study_id=phs000421.v1.p1 through dbGaP accession number phs000421.v1.p1. The grants that have funded the enrollment of the cases and controls to be used in this GWAS are: R01EY016514 (DUEC, PI: Gordon Klintworth), R01EY016482 (CWRU, PI: Sudha Iyengar), and 1X01HG006619-01 (PI: Sudha Iyengar, Natalie Afshari). We would like to thank the FECD participants and the FECD Research Group for their valuable contribution to this research.

The dataset(s) used for the analyses described in this manuscript were obtained from the Age-Related Eye Disease Study (AREDS) Database found at https://www.ncbi.nlm.nih.gov/projects/gap/cgi-bin/study.cgi?study_id=phs000429.v1.p1 through dbGaP accession number phs000001.v3.p1. Funding support for AREDS was provided by the National Eye Institute (N01-EY-0-2127). We would like to thank the AREDS participants and the AREDS Research Group for their valuable contribution to this research.

The dataset(s) used for the analyses described in this manuscript were obtained from the NEI Study of Age-Related Macular Degeneration (NEI-AMD) Database found at https://www.ncbi.nlm.nih.gov/projects/gap/cgi-bin/study.cgi?study_id=phs000246.v2.p1 through dbGaP accession number phs000684.v1.p1. Funding support for NEI-AMD was provided by the National Eye Institute. We would like to thank NEI-AMD participants and the NEI-AMD Research Group for their valuable contribution to this research.

